# A wind speed threshold for increased outdoor transmission of coronavirus: An ecological study

**DOI:** 10.1101/2021.02.05.21251179

**Authors:** Sean A. P. Clouston, Olga Morozova, Jaymie R. Meliker

## Abstract

**Background:** To examine whether outdoor transmission may contribute to the COVID-19 epidemic, we hypothesized that slower outdoor wind speed is associated with increased risk of transmission when individuals socialize outside.

**Methods:** Daily COVID-19 incidence reported in Suffolk County, NY, between March 16^th^ – December 31^st^, 2020, was the outcome. Average wind speed and maximal daily temperature were collated by the National Oceanic and Atmospheric Administration. Negative binomial regression was used to model incidence rates while adjusting for susceptible population size.

**Results:** Cases were very high in the initial wave but diminished once lockdown procedures were enacted. Most days between May 1^st^, 2020, and October 24^th^, 2020, had temperatures 16-28°C and wind speed diminished slowly over the year and began to increase again in December 2020. Unadjusted and multivariable-adjusted analyses revealed that days with temperatures ranging between 16-28°C where wind speed was <8.85 kilometers per hour (KPH) had increased COVID-19 incidence (aIRR=1.45, 95% C.I.=[1.28-1.64], P<0.001) as compared to days with average wind speed ≥8.85 KPH.

**Conclusion:** Throughout the U.S. epidemic, the role of outdoor shared spaces such as parks and beaches has been a topic of considerable interest. This study suggests that outdoor transmission of COVID-19 may occur by noting that the risk of transmission of COVID-19 in the summer was higher on days with low wind speed. Outdoor use of increased physical distance between individuals, improved air circulation, and use of masks may be helpful in some outdoor environments where airflow is limited.

## Background

The novel severe acute respiratory syndrome coronavirus-2 (SARS-CoV-2) that causes a potentially deadly disease called coronavirus disease 2019 (COVID-19), began spreading in China [1], and Italy [2] before arriving in the United States (U.S.). COVID-19 first hit in the U.S. in regions, such as New York (N.Y.) and California, where global travelers often arrive into the U.S. [3]. Suffolk County, N.Y., experienced its first wave of infections early in March 2020, when the pandemic had just arrived in N.Y., causing a high degree of transmission and large numbers of COVID-related deaths.

COVID-19 transmits *via* aerosolized viral particles that begin shedding before symptoms are evident [4], making it difficult to trace patterns or locations where exposures are occurring. As a result, approximately half of those diagnosed with COVID-19 report not knowing where they may have become infected [5]. One explanation for a lack of known exposures is that COVID-19 transmits in spaces that are believed to be safe. A handful of studies have made some headway in identifying such situations. For example, one study found that COVID-19 could transmit through the air over relatively long distances [6] and another highlighted the impact of air conditioning vents [7]. A third study found that a cluster of 17 cases were traced to indirect transmission in shared spaces at a shopping mall in Wenzhou, China [8]. Still other studies have revealed that individuals in a constricted space could spread COVID-19 *via* inhaled transmission over potentially large distances by following airflow within a restaurant [7] and the Diamond Princess cruise ship [6].

A recent review concluded that transmission within constricted indoor spaces is critically important [9]. However, outdoor exposures have been reported, yet relatively little is known about conditions that reduce safety of outdoor social contacts. There are reports of sporadic outbreaks in outdoor environments, including at a construction site in Singapore [10, 11], jogging [10], or during conversation [12]. Because of much lower risk outdoors [13], close outdoor contacts are often described as being risk-free, and exposure-mitigating strategies have focused on promoting the use of exterior spaces when conducting social activities in efforts to mitigate risk of exposure. Outdoor gatherings (for example, participating in events such as backyard barbecues, sitting near to others while watching outdoor events, standing in line outside, or socializing outdoors), may be sensitive to circumstances that may influence their protective features. If exposure occurs outside, simulation studies suggest that transmission may be hampered by the same factors as are commonly seen in studies of indoor transmission including the air turnover rate [13]. Indeed, one preliminary study reported evidence of an association globally between weather dynamics including lower temperatures and lower wind speed with small increase in COVID-19 incidence [14] that may be non-linear [15], with at least one locality in Indonesia reporting local findings supporting this pathway [16].

The present study examined data reported in Suffolk County, N.Y., a large suburban county (∼1.5 million) that reported 96,057 cases between March and December 2020. The existence of a non-linearity in associations could imply that wind speed is moderated by another factor, potentially human activity. In the present study, we hypothesized that lower exterior wind speed would be associated with an increased risk of transmission during days ranging in temperature from 16-28°C (degrees Celsius, equivalent to 60-84 degrees Fahrenheit [°F]) when individuals were most likely to be socializing outside.

## Methods

### Setting

Suffolk County is a large county (2,362 square-kilometers [km^2^]) of approximately 1.5-million people that predominantly acts as an exterior suburban community serving New York City. The median age is 41.8 years; 66.6% are non-Hispanic White, 20.2% are Hispanic, 8.8% are Black, while the remainder predominantly reports being Asian or having two or more races. The median household income in Suffolk County is 54.6% higher than the national average. Overall, 6.8% of households fall below the national poverty line and 5.2% report lacking health insurance. Suffolk County is relatively densely populated with 645.6 people/km^2^.

### Measures

To examine the potential for exterior exposure risk, we modeled COVID-19 incidence using cases reported to the Suffolk County Department of Health from March 16^th^, when data first began being recorded reliably using an electronic interface, until December 31^st^, 2020. At that time, Suffolk County was enduring a second wave. Daily case counts were shared with Stony Brook University to support the COVID-19 modeling efforts at the local level. After cleaning, county-level data were published online to a publicly-accessible database (the Supplemental Data Resource provides cleaned county-level data merged with other variables used in this study). We limited the analysis to dates following March 16^th^, 2020, with the opening of multiple drive-through testing sites throughout the area and the establishment of regular case-reporting routines. Susceptible population estimates integrate overall county residential estimates derived from the U.S. census and were updated for daily death counts, and for the reported number of COVID-19-related disease counts.

Since daily case counts exhibit temporal dependence that is primarily determined by the unobserved community force of infection, in secondary analyses we examined an alternative outcome measure of relative change in daily case counts compared to an 8-day forward/backward autoregressive moving average [17], as defined by:

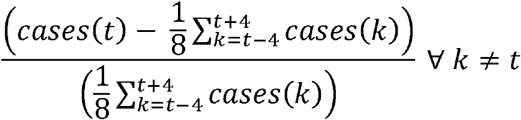

The 8-day forward/backward moving average, when integrated into the model, serves as a proxy measure of underlying force of infection. This allows us to partially capture the variability in absolute case counts that is due to “natural” transmission patterns rather than external shocks such as wind speed. It is important to note that, on average, this measure would be zero when case counts remain relatively constant over time, however, this measure will track the periods of exponential rise (where it will be positive) and decay (where it will be negative) of an epidemic’s waves. It is therefore important to take these distinct behaviors into account.

*Maximal daily temperature*, as well as *average wind speed*, were derived from the U.S. *National Oceanic and Atmospheric Administration* data portal (w2.weather.gov). Data were recorded at a central location at the MacArthur Airport in Islip, N.Y. Total snowfall and rainfall were recorded in inches and converted to centimeters. While temperatures 16-28°C are likely to be protective, reduced wind speed impact on these days may emerge because individuals are more likely to be socializing outdoors where risk is markedly lower. In the summer, higher wind speed increases airflow and may reduce risk *versus* in the winter when it may work to push outside social contacts to shelter in indoor spaces. When exterior temperatures are warm enough (16-28°C) to allow for outdoor social contacts to occur comfortably, we anticipated that increased wind speed would reduce overall transmission risk. In contrast, on days where exterior temperatures were cooler, increased wind speed might cause individuals to retreat indoors for social occasions.

### Covariates

We adjusted for the number of days since lockdown (March 16^th^, 2020) and days since reopening began (May 15^th^, 2020) in Suffolk County, N.Y. To account for differences in daily reporting patterns, we incorporated a categorical variable indicating the day of the week that cases were reported. Noting that there have been significant spread following holidays, we incorporated an indicator of holidays that also incorporated the most significant weekend nearby. We also included covariates measuring rainfall and snowfall because they may correlate with wind speed as well as social activities outdoors. In the primary analysis, we also adjusted for the 8-day forward/backward moving average daily case count.

### Statistical Modeling

Descriptive characteristics include time-related trends in maximal temperature, average daily wind speed, and daily case counts. Daily and smoothed trends in maximal temperature and in average wind speed were reported.

In the main analysis, the incidence of COVID-19 positive caseload was reported as case counts per day so multivariable-adjusted modeling relied on negative binomial regression [18]. Negative binomial regression was chosen over alternatives including Poisson because we were concerned about the potential for over-dispersion in the outcome [19] since the infectious disease caseload is highly variable and because COVID-19 appears to spread commonly through super-spreading clusters [20]. A nine-day lag between exposure and case registration was assumed, consistent with epidemiological estimates of the incubation period for COVID-19 [21, 22] coupled with a two-day testing and one-day reporting lag period that has been common in Suffolk County since testing became widely available. Unadjusted and multivariable-adjusted incidence rate ratios (IRR) and 95% confidence intervals (95% C.I.) were reported. The interval between infection and disease ascertainment is unobserved and varies geographically by local testing availability and reporting systems: it can be reduced in places where testing is easy to find and lengthened in places where testing is difficult or requires hospitalization. As such, we conduct a sensitivity analysis considering the range of values of time intervals between exposure and case reporting. For our lagging period, we allowed four days because our experience suggests that it takes two days to report testing results to the Department of Health, and an additional day to report those results publicly. Fifteen days was selected as a ceiling for index case analysis to reduce the risk of sequential outcomes from prior case/exposure cycles consistent with prior publications [23]. However, in sensitivity analyses we report results for a 4–13-day range to clarify the impact of those choices. We used the log-likelihood to compare model fit for different lags.

We analyzed the secondary outcome – a relative measure of daily case counts calculated as ln(incident cases/population * 100,000) – using linear regression with the same set of covariates as the primary outcome measure and exploring the results for a range of reporting lags.

Since we theorized that there is heterogeneity in association between wind speed and COVID-19 transmission may depending on temperature, cutoffs for “warm” days and for days when wind speed was sufficiently fast were determined by comparing Akaike’s information criterion (AIC) across multiple models using different details as modeled parameters. We compared AIC between models to determine that 16°C (60 °F) was an optimal lower bound in temperature, while follow-up analyses revealed an upper bound of 28°C (84°F). To account for seasonality, we also adjusted for the maximal daily temperature. Because cutoffs may be useful when adjudicating risk at the local level, we used AIC to identify optimal cutoffs for wind speed. This resulted in identifying *low* wind speed to be <8.85 KPH (kilometers per hour (KPH), equivalent to approximately 5.5 miles per hour).

Since the relative measure of daily case counts only partially adjusts for the community force of infection and underlying “natural” epidemic dynamics, we also conducted additional stratified sensitivity analyses cut into periods when case counts were relatively flat (06/07/2020-11/03/2020) and when the epidemic was exponentially increasing (03/16/2020-04/10/2020 and 11/04/2020-12/31/2020) or decaying (04/11/2020-06/06/2020). We used two criteria: daily temperature and epidemic dynamics pattern (flat *versus* rising/falling) to determine subsets for stratified analyses. Analyses were completed using Stata 16/MP [StataCorp].

## Results

We begin by showing the number of daily cases over the entire observational window (Figure 1). Cases were very high in the initial wave but diminished quickly once lockdown procedures were enacted.

**Figure 1.**
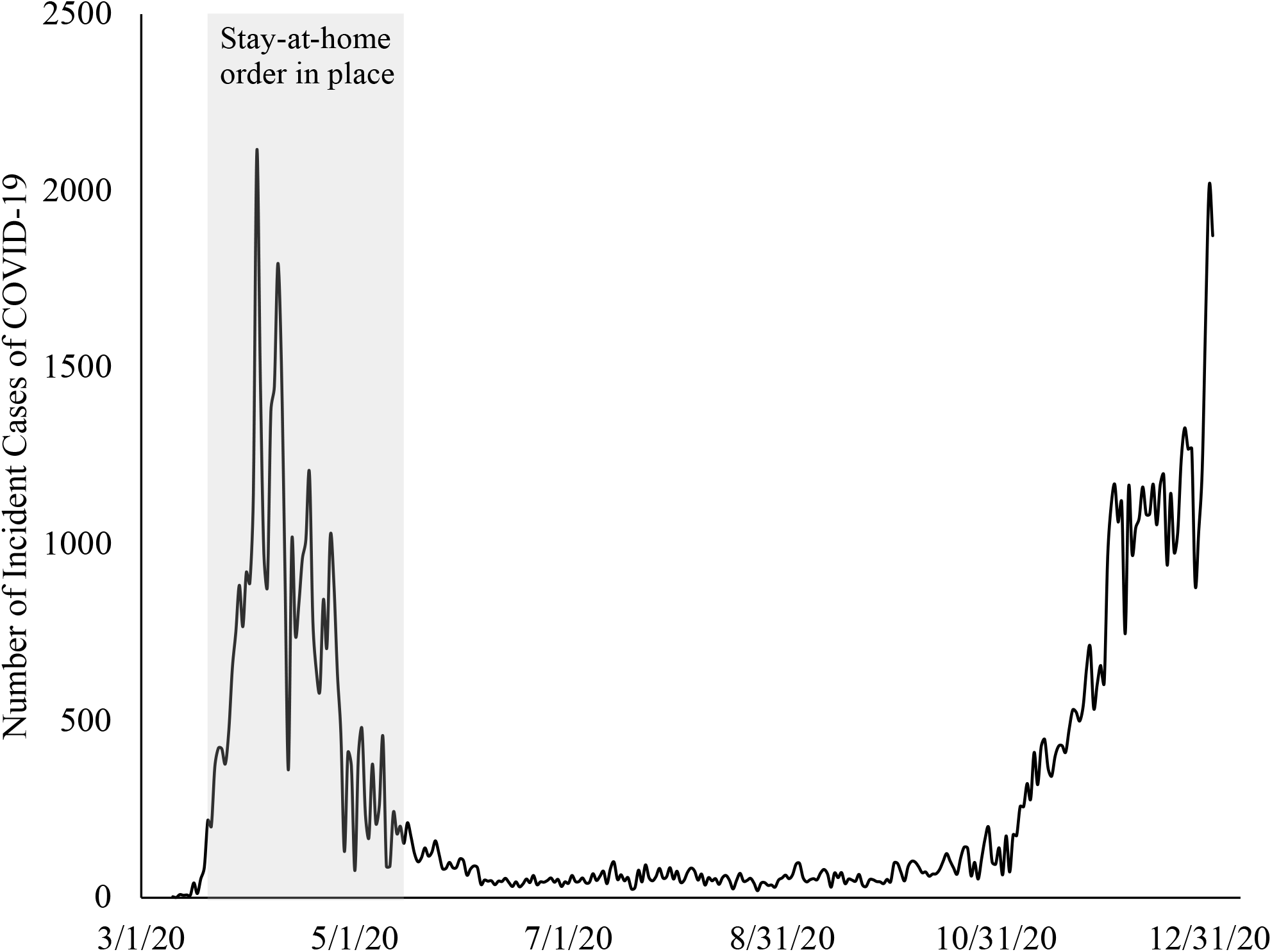
Trends in daily COVID-19 cases identified in Suffolk County from March 16^th^ – December 31^st^, 2020.

The average temperature was 19.8 ± 8°C and the average daily wind speed was 14.0 ± 5.8 KPH. Trends in daily temperature and wind speed are depicted throughout the analytic period (Figure 2). Most days between May 1^st^, 2020, and October 24^th^, 2020, were characterized by temperatures 16-28°C (solid red lines show this range). The trend in average wind speed (black dashed line) diminished slowly over time and then began to increase again in December 2020.

**Figure 2.**
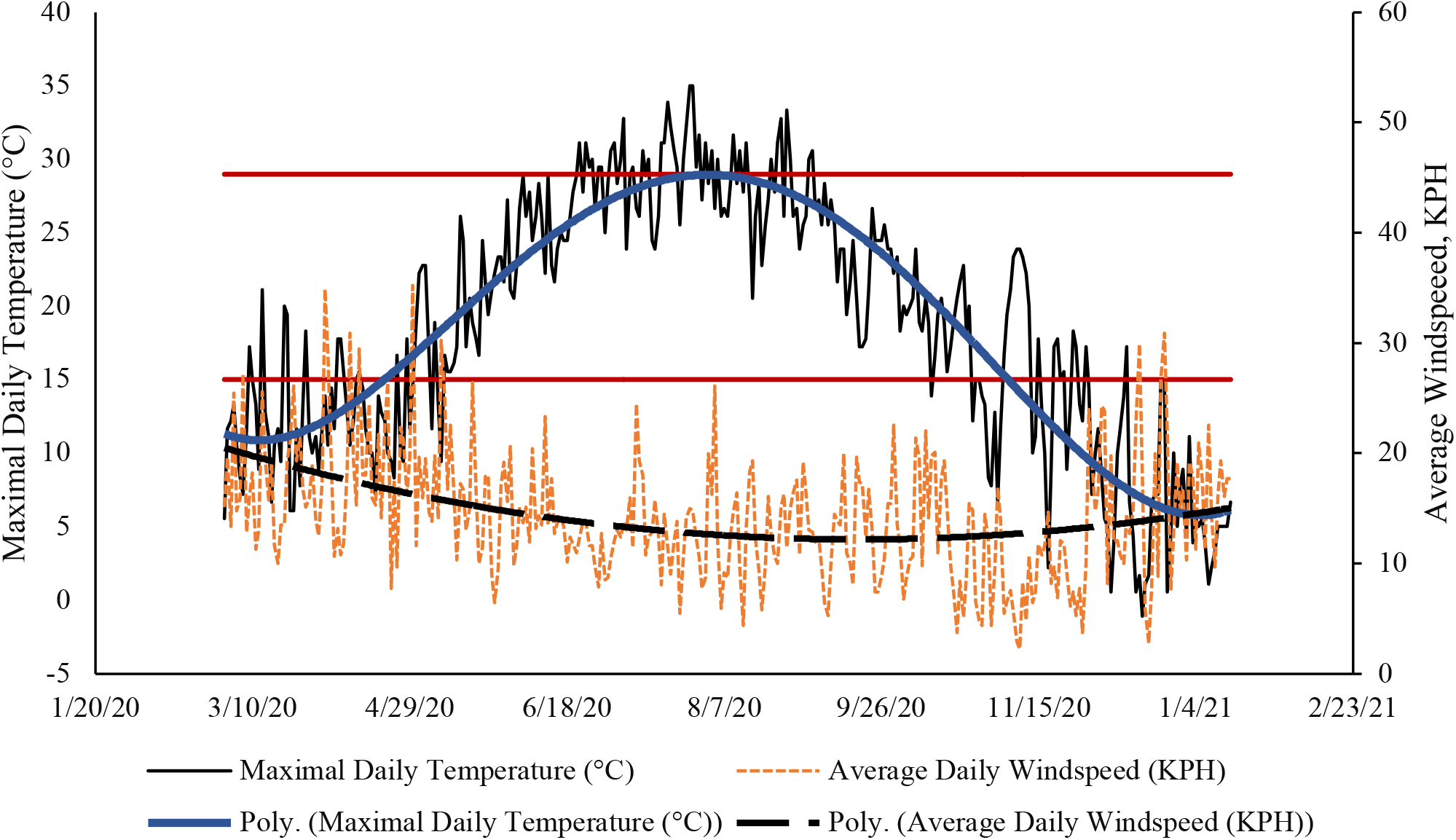
Trends in maximal daily temperatures, expressed in °C, and mean daily wind speed expressed in kilometers per hour in Suffolk County, NY, from March 16^th^ – December 31^st^, 2020. The horizontal red lines show temperatures in the 16-28°C range.

Further interrogating the functional shape of the relationship between the wind speed and incidence of COVID-19 (Figure 3) we found that during periods where temperatures ranged from 16-28°C, reduced wind speed was associated with increased incidence. However, on cooler days, when very high wind speeds were most common, incidence of COVID-19 appears to increase slightly as a function of wind speed though this was not evident in multivariable analyses. Using the logarithmic transformation to capture tapering threshold effects in a multivariable-adjusted model examining the impact of wind speed only on days that were 16-28°C. Exploring the implications of this threshold effect we found that while an increase in wind speed from 5 to 6 KPH was associated with a 12.56% caseload reduction, a similar increase from 15 to 16 KPH was only associated with a 1.16% decrease in caseload. Visual inspection showed that on warm days (temperatures ranging from 16-28°C) with very high wind speed (above 20 KPH) increasing wind speed was associated with increased transmission. However, using a quadratic transformation we did not find this association to be statistically significant (P = 0.071).

**Figure 3.**
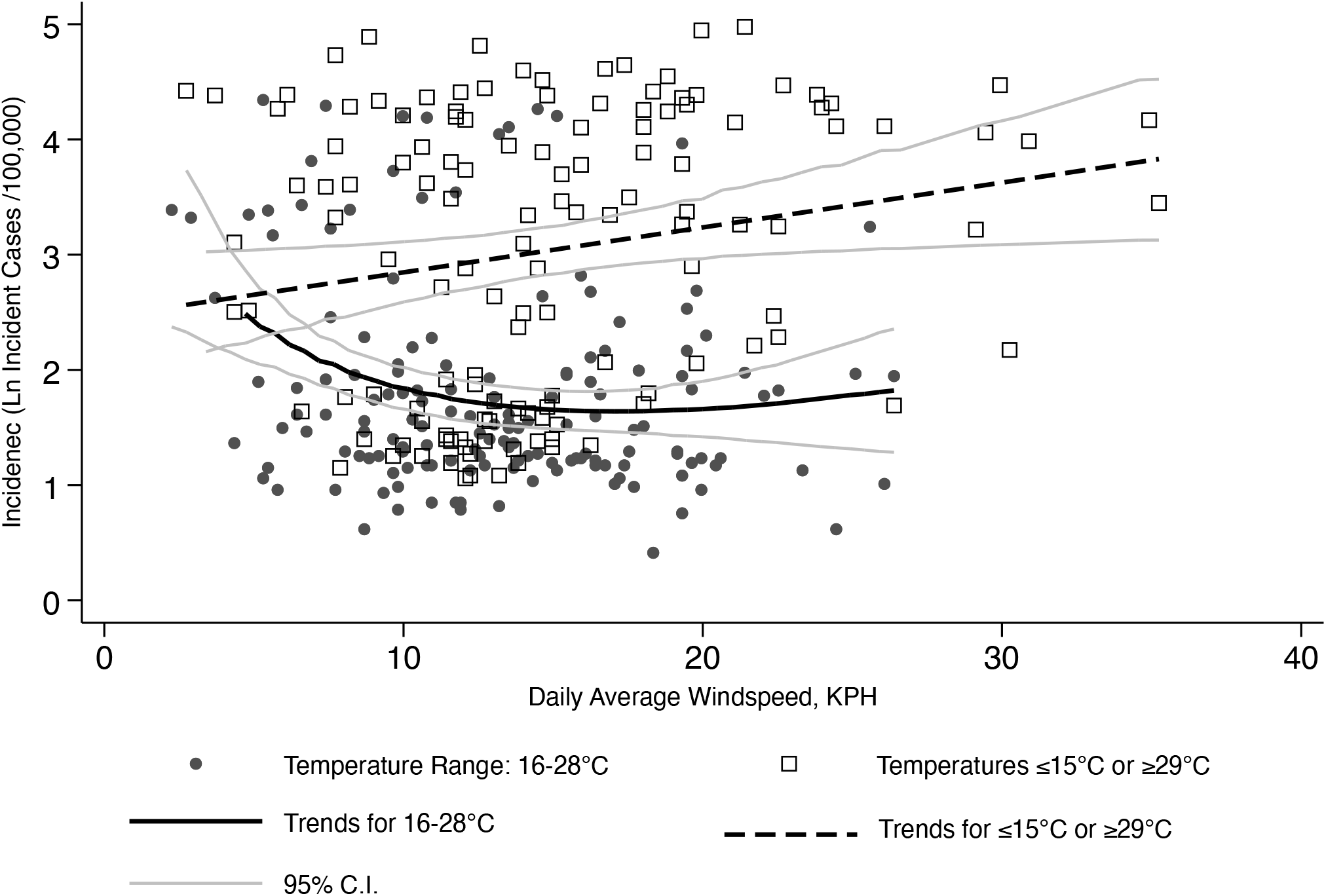
Average wind speed versus number of incident cases of COVID-19 in Suffolk County from March 16^th^ – December 31^st^, 2020 **Note:** The natural log function was selected because it performed better (AIC = 4055.4) than alternative specifications including linear (AIC = 4105.3), inverse (AIC = 4298.5), and quadratic (AIC = 4057.3). Unadjusted and multivariable-adjusted models are shown in Table 2. Note that the incidence of COVID-19 was lagged from wind speeds by nine days.

Unadjusted analyses revealed statistically significant associations between higher COVID-19 incidence and lower wind speed in 16-28°C weather (**Table 1**). Multivariable-adjusted analyses similarly revealed that results remained statistically significant upon adjusting for confounders.

**Table 1.** Incidence rate ratios for COVID-19 derived from negative binomial regression showing both unadjusted and multivariable-adjusted analyses from March 16^th^ – December 31^st^, 2020

| Variable | Unadjusted | Multivariable-Adjusted |
| --- | --- | --- |
|  | IRR [95% C.I.] | aIRR [95% C.I.] |
| Wind speed (Ln – KPH) when temperature 16-28°C | 1.85 [1.67-2.06]<br>P < 0.001 | 1.17 [1.09-1.26]<br>P < 0.001 |
| Wind speed (Ln - KPH) when temperature ≤15 or ≥29°C | 1.04 [0.79-1.36]<br>P = 0.798 | 0.93 [0.81-1.06]<br>P = 0.248 |
| Maximal Exterior Temperature, °C |  | 1.00 [0.99-1.01]<br>P = 0.677 |
| Days since Lockdown |  | 0.95 [0.93-0.97]<br>P < 0.001 |
| Days since Reopening |  | 1.06 [1.04-1.08]<br>P < 0.001 |
| Holiday Adjustment |  | 1.11 [0.92-1.33]<br>P = 0.264 |
| Snowfall, cm |  | 0.98 [0.92-1.04]<br>P = 0.540 |
| Rainfall, cm |  | 1.01 [0.94-1.09]<br>1.02 P = 0.776 |
| Eight-day forward/backward moving average |  | 1.22 [1.20-1.25]<br>P < 0.001 |
| $\alpha$ | 0.98 [0.85-1.13] | 0.17 [0.14-0.2] |

**Note:** IRR: incidence rate ratio; 95% C.I.: 95% confidence interval. All models additionally adjust for day of the week in which cases were reported and for the size of the county population adjusted for reductions due to death or recovery from COVID-19 during the period of observation. $\alpha$ is a measure of dispersion. P-values derived from Student's T-tests.

As noted in the Methods section, optimal temperature cutoffs were 16-28°C in temperature, and <8.85 KPH in wind speed. Using these cutoffs, in Table 2 we examined the risk associated with lower wind speed (<8.85 KPH) on days with maximal temperatures in the 16-28°C range. Analyses revealed that on days with temperatures from 16-28°C, exposures to wind speed <8.85 KPH was associated with a 45% increase in incidence in multivariable-adjusted models.

**Table 2.** Incidence rate ratios for COVID-19 derived from negative binomial regression showing both unadjusted and multivariable-adjusted analyses comparing days where wind speed <8.85 KPH to days with ≥8.85 KPH wind speeds from March 16^th^ – December 31^st^, 2020

| Variable | Unadjusted<br>IRR [95% C.I.] | Multivariable Adjusted<br>aIRR [95% C.I.] |
| --- | --- | --- |
| Wind speed <8.85 KPH when temperature 16-28°C | 4.09 [3.16-5.28]<br>P < 0.001 | 1.45 [1.28-1.64]<br>P < 0.001 |
| Wind speed <8.85 KPH when temperature $\leq 15$ or $\geq 29$°C | 1.45 [1.04-2.03]<br>P = 0.029 | 1.03 [0.88-1.20]<br>P = 0.717 |
| Maximal Exterior Temperature, °C |  | 0.99 [0.99-1.00]<br>P = 0.021 |
| Days since Lockdown |  | 0.94 [0.93-0.96]<br>P < 0.001 |
| Days since Reopening |  | 1.07 [1.05-1.09]<br>P < 0.001 |
| Holiday Adjustment |  | 1.12 [0.94-1.34]<br>P = 0.208 |
| Snowfall, mm |  | 0.93 [0.82-1.06]<br>P = 0.274 |
| Rainfall, mm |  | 1.02 [0.88-1.20]<br>P = 0.774 |
| Eight-day forward/backward moving average |  | 1.21 [1.19-1.24]<br>P < 0.001 |
| $\alpha$ | 1.02 [0.88-1.17] | 0.16 [0.13-0.19] |

**Note:** \* KPH: Kilometers Per Hour; °C: degrees Celsius; IRR: incidence rate ratio; 95% C.I.: 95% confidence interval. All models adjust for day of the week in which cases were reported and for the size of the county population adjusted for reductions due to individuals who had died or become immune due to COVID-19 during the period of observation. $\alpha$ is a measure of dispersion. P-values derived from Student's T test.

### Sensitivity Analysis

We examined the sensitivity of the results to analytic choices by first examining whether reliance on different outcomes made differences to the results. For the relative change in daily case counts compared to an 8-day forward/backward moving average, the results were substantively similar (B = -16.12 [-27.78, -4.45], P=0.007) on days with temperatures from 16-28°C; in other words, days where wind speed was <8.85 KPH were attributed with 16.12% increases in relative incidence (Supplemental Table 1). We also examined whether choices in the lag between exposure and case reporting changed our results. While the results shown theoretically represent the appropriate timing, we also examined variation in periods between exposure and case recording from 4-13 days. We found that while the nine-day reporting average was the best performing within our hypothesized observational window (Supplemental Figure 1). Across all lags, we identified a consistent association between slower wind speed days and lower follow-up case counts (Supplemental Table 2). We examined whether holidays were more impactful depending on temperature but found that while the effect sizes were slightly smaller on days with temperatures from 16-28°C (interaction B = -0.18, P=0.142), these differences were not statistically significant. Finally, we stratified analysis dates into periods characterized by rising, falling, and stable transmission. This analysis resulted in the same overall association (aIRR_>8.85 KPH_ = 0.87 [0.75-1.03]; aIRR_Ln-KPH_ = 0.88 [0.75-1.05]) though insufficient observations to achieve statistical power (power = 0.65).

## Discussion

The COVID-19 pandemic has caused an immense toll on the American population and has inflicted enormous economic damage. Current evidence suggests that COVID-19 is airborne and is predominantly spread indoors. The present study examined variations in wind speed under the hypothesis that higher winds may disperse COVID-19 viral particles away from people socializing outdoors, thereby offering increased protection among individuals who may have been exposed to COVID-19 outdoors. We found that slow average wind speed (<8.85 KPH) was associated with increased incidence of COVID-19 on days that had temperatures supporting socializing outdoors (16-28°C; aIRR = 1.45 [1.28-1.64], P < 0.001). This study supports the view that while transmission was lowest when days were in comfortable ranges (from 16-28°C), on these days the risk was highest when wind speed was slow.

This study suggests that low wind speed may reduce the protective impact of weather ranging from 16-28°C. Results align with anecdotal reports from local Departments of Health and from the Centers for Disease Control and Prevention [24], who have noted that gatherings of increased risk include outdoor social gatherings such as “Backyard Barbecues”. One interpretation of this evidence may be that airborne transmission in shared outdoor spaces is feasible on days when the wind is insufficient to disperse viral particles. For example, wind speed in weather outside of the 16-28°C temperate zone may make social activities less pleasant or may increase the risk of transmission in outdoor settings with stale air.

The present study represents a step forward to understanding the regional role of outdoor wind and temperature dynamics, and their interrelationships when trying to understand COVID-19 infection dynamics. The next steps in this research area might include the study of microclimate dynamics within regions to determine the relevance of architectural design, fencing, and wind flow within roads in determining geographical differences in disease transmission and exposure dynamics. Understanding the geographical distribution of cases resulting on wind-less compared to similar windy days may help determine other factors, such as population density or housing density, that modify impact of reduced wind speed. Additionally, multilevel analyses might examine the extent to which social activities might be affected most by reduced wind speed. Yet, while geographic targets are critical, further research is also needed to determine the extent to which reduced wind speed is more, or less, impactful with novel COVID variants or with other respiratory diseases. One potential output of such information may be to inform the creation of a weather warning system so that individuals or policymakers could issue guidelines or warning systems when masking usage might be recommended outdoors or in outdoor spaces at risk of reduced wind speed.

### Limitations

Despite examining a large population (∼1.5 million) that identified many cases (96,057 between March-December 2020), this study is limited in examining the experience of a single U.S. County. Although there is little reason to think that shared indoor spaces would increase on days of lower wind speed in the 16-28°C temperature range, we cannot conclusively state that higher wind speed protected any individuals. Our results were strongly influenced by covariates as evidenced by the change in IRR observed in unadjusted *versus* adjusted models; it is always possible that key confounders were missing from our model. For example, we could not address the potential for non-independence that may emerge when individuals who have previously survived COVID-19 may be re-infected. However, sensitivity analyses examining the percent change of new cases on a given day relative to the eight-day backward/forward average case count attempted to address temporal changes in incidence patterns directly within the outcome variable, and our results were similar. Follow-up research is necessary to determine specifics about exposures, including distances that COVID-19 viral particles can travel and reliably infect individuals and microclimate differences that may affect specific geographic differences that may moderate these results.

To obtain a measure of wind speed for this analysis, we relied on data from a central airport. While this provided consistent measures of wind speed across the island, these measures may not be generalizable to microclimates occurring in the fenced-in backyards, lea of hills and dunes, or forests. Notably, this choice may mean that cutoffs used here may not apply in other situations. More analysis is necessary if weather data are going to be relied upon to help understand caseload in other areas. We reported a nine-day exposure-test positive reporting lag structure; however, sensitivity analyses suggested that a 16-day lag structure may work better. The 16-day lag is outside of the expected lag period for cases in our area. Still, we felt that it might indicate that case dynamics could proceed from asymptomatic younger individuals to cause secondary cases in older individuals reported 16 days later. As such, future work should anticipate that different cutoffs will be necessary when wind speeds are measured in other places and in locations where wind is highly sensitive to local geography.

## Conclusions

Throughout the U.S. epidemic, the role of outdoor shared spaces such as parks and beaches has been studied, and ultimately beaches and parks remained open because outdoor gatherings are considerably less risky than indoor ones. This analysis does little to suggest that either should be closed, since the level of risk due to outdoor exposures should be weighed in relation to the much higher risk of exposure in shared interior spaces such as houses, restaurants, or public transport. Instead, this study may suggest that individuals socializing outdoors may not be completely safe by being outdoors and should remain vigilant, especially on days where airborne particles may be less likely to disperse due to contextual factors such as reduced wind speed, that may reduce the benefits of socializing outside. In this case, outdoor use of increased physical distance between individuals, improved air circulation, and use of masks may be helpful in some outdoor environments where airflow is limited.

## Data Availability

Data can be retrieved from multiple sources. Data will be made available to researchers upon publication of the manuscript.

## Ethics approval and consent to participate

Data used in this study are secondary analyses of daily case counts reported on publicly available websites, and therefore was not human subject’s research. All identifiers were removed from the data prior to publication online. No administrative privileges were needed to access the data used in this study.

## Consent for publication

Not Applicable.

## Availability of data and material

All data analyzed during this study are included in this published article and its supplementary information files.

## Competing interests

None.

## Funding

None.

## Authors’ contributions

SC analyzed data and drafted the manuscript. OM provided scientific oversight, edited the manuscript, and provided topical expertise. JM provided scientific oversight and critically edited the manuscript.

## Acknowledgements

None.

## Notes

### Competing Interest Statement

The authors have declared no competing interest.

### Author Declarations

The analysis of publicly available deidentified case counts retrieved from the internet are considered to be not human subjects research and are exempt from ethics review.

### Summary of Updates

We are updating with the post peer-review manuscript.

## REFERENCES

1. Novel CPERE: The epidemiological characteristics of an outbreak of 2019 novel coronavirus diseases (COVID-19) in China. Zhonghua liu xing bing xue za zhi= Zhonghua liuxingbingxue zazhi 2020, 41(2):145.

2. Cereda D, Tirani M, Rovida F, Demicheli V, Ajelli M, Poletti P, Trentini F, Guzzetta G, Marziano V, Barone A: The early phase of the COVID-19 outbreak in Lombardy, Italy. In.: Arxiv; 2020.

3. Clouston SA, Natale G, Link B: Socioeconomic inequalities in the spread of coronavirus-19 in the United States: A examination of the emergence of social inequalities. Social Science & Medicine 2020:113554.

4. Morawska L, Milton DK: It Is Time to Address Airborne Transmission of Coronavirus Disease 2019 (COVID-19). Clinical Infectious Diseases 2020, 71(9):2311–2313.

5. Tenforde MW, Rose EB, Lindsell CJ, Shapiro NI, Files DC, Gibbs KW, Prekker ME, Steingrub JS, Smithline HA, Gong MN: Characteristics of adult outpatients and inpatients with COVID-19—11 academic medical centers, United States, March– May 2020. Morbidity and Mortality Weekly Report 2020, 69(26):841.

6. Almilaji O, Thomas P: Air recirculation role in the infection with COVID-19, lessons learned from Diamond Princess cruise ship. medRxiv 2020.

7. Lu J, Gu J, Li K, Xu C, Su W, Lai Z, Zhou D, Yu C, Xu B, Yang Z: COVID-19 outbreak associated with air conditioning in restaurant, Guangzhou, China, 2020. Emerging infectious diseases 2020, 26(7):1628.

8. Cai J, Sun W, Huang J, Gamber M, Wu J, He G: Indirect virus transmission in cluster of COVID-19 cases, Wenzhou, China, 2020. 2020.

9. Bulfone TC, Malekinejad M, Rutherford GW, Razani N: Outdoor Transmission of SARS-CoV-2 and Other Respiratory Viruses, a Systematic Review. The Journal of Infectious Diseases 2020.

10. Leclerc QJ, Fuller NM, Knight LE, Funk S, Knight GM, Group CC-W: What settings have been linked to SARS-CoV-2 transmission clusters? Wellcome Open Research 2020, 5(83):83.

11. Lan F-Y, Wei C-F, Hsu Y-T, Christiani DC, Kales SN: Work-related COVID-19 transmission in six Asian countries/areas: A follow-up study. PloS one 2020, 15(5):e0233588.

12. Qian H, Miao T, Li L, Zheng X, Luo D, Li Y: Indoor transmission of SARS-CoV-2. medRxiv 2020.

13. Rowe BR, Canosa A, Drouffe JM, Mitchell JBA: Simple quantitative assessment of the outdoor versus indoor airborne transmission of viruses and COVID-19. Environmental Research 2021, 198:111189.

14. Islam N, Shabnam S, Erzurumluoglu AM: Temperature, humidity, and wind speed are associated with lower Covid-19 incidence. MedRxiv 2020.

15. Yuan J, Wu Y, Jing W, Liu J, Du M, Wang Y, Liu M: Non-linear correlation between daily new cases of COVID-19 and meteorological factors in 127 countries. Environmental Research 2021, 193:110521.

16. Rendana M: Impact of the wind conditions on COVID-19 pandemic: A new insight for direction of the spread of the virus. Urban climate 2020, 34:100680.

17. Ozaki T: On the order determination of ARIMA models. Journal of the Royal Statistical Society: Series C (Applied Statistics) 1977, 26(3):290–301.

18. Long JS, Freese J: Regression models for categorical dependent variables using Stata: Stata press; 2006.

19. Gardner W, Mulvey EP, Shaw EC: Regression analyses of counts and rates: Poisson, overdispersed Poisson, and negative binomial models. Psychological bulletin 1995, 118(3):392.

20. Wong F, Collins JJ: Evidence that coronavirus superspreading is fat-tailed. Proceedings of the National Academy of Sciences 2020, 117(47):29416–29418.

21. Biggerstaff M, Cowling BJ, Cucunubá ZM, Dinh L, Ferguson NM, Gao H, Hill V, Imai N, Johansson MA, Kada S: Early insights from statistical and mathematical modeling of key epidemiologic parameters of COVID-19. Emerging infectious diseases 2020, 26(11).

22. McAloon C, Collins Á, Hunt K, Barber A, Byrne AW, Butler F, Casey M, Griffin J, Lane E, McEvoy D: Incubation period of COVID-19: a rapid systematic review and meta-analysis of observational research. BMJ open 2020, 10(8):e039652.

23. Xi W, Pei T, Liu Q, Song C, Liu Y, Chen X, Ma J, Zhang Z: Quantifying the time-lag effects of human mobility on the COVID-19 transmission: A Multi-City Study in China. Ieee Access 2020, 8:216752–216761.

24. Tanne JH: Covid-19: Cases still rising in at least 23 US states as health officials warn against gatherings. bmj 2020, 369.

