## Supplemental Tables and Figures for "A wind speed threshold for increased outdoor transmission of coronavirus: An ecological study"

**Supplemental Table 1.** Multivariable-adjusted regression coefficient, 95% confidence interval, and P-values examining association between wind speed (<8.85 KPH) on days with temperatures of 16-28°C and percentage increases in case numbers

| Variable | Coef., 95% C.I., P |
| --- | --- |
| Wind speed <8.85 KPH when temperature 16-28°C | -16.12 [-4.45, -27.78] P =0.007 |
| Wind speed <8.85 KPH when temperature ≤15 or ≥29°C | -0.80 [-3.03, 1.43] P =0.479 |
| Maximal Exterior Temperature, °C | 0.06 [-1.09, 1.22] P =0.913 |
| Lockdown | -0.56 [-2.73, 1.61] P =0.611 |
| Reopening | 0.61 [-1.79, 3.02] P =0.615 |
| Holiday Adjustment | 8.87 [-16.41, 34.15] P =0.490 |
| Snowfall, cm | 0.40 [-19.24, 20.05] P =0.968 |
| Rainfall, cm | -0.85 [-22.25, 20.55] P =0.938 |
| Eight-day forward/backward moving average | -2.26 [-4.80, 0.28] P =0.081 |

**Supplemental Table 2.** Multivariable-adjusted incidence rate ratio (aIRR), 95% confidence interval, and P-values examining association between wind speed (Ln-KPH) on days with temperatures from 16-28°C and increases in case numbers only in samples where a 13-day lag was observed to maintain predictive stability.

| Lagged Days | aIRR | 95% C.I. | P |
| --- | --- | --- | --- |
| 5 | 1.154 | 1.053-1.264 | 0.002 |
| 6 | 1.132 | 1.033-1.241 | 0.008 |
| 7 | 1.060 | 0.967-1.161 | 0.214 |
| 8 | 1.124 | 1.027-1.231 | 0.011 |
| **9** | 1.205 | 1.102-1.318 | <0.001 |
| 10 | 1.147 | 1.048-1.256 | 0.003 |
| 11 | 1.085 | 0.993-1.186 | 0.072 |
| 12 | 1.153 | 1.058-1.256 | 0.001 |
| 13 | 1.110 | 1.018-1.21 | 0.019 |

**Supplemental Figure 1.** Gaussian-smoothed fit characteristics for the model presented in Table 1 relying on different lag structures examining possible lags of 4-13 days. Note that the best fitting lag (9 days) was shown using a red diamond.

| Lagged Days | Log Likeliihood |
| --- | --- |
| 5 | -1635.47 |
| 6 | -1634.63 |
| 7 | -1633.56 |
| 8 | -1632.03 |
| 9 | -1630.9 |
| 10 | -1631.32 |
| 11 | -1632.21 |
| 12 | -1632.54 |
| 13 | -1632.62 |
| 14 | -1631.33 |
| 15 | -1628.21 |
| 16 | -1626.33 |
| 17 | -1628.02 |
| 18 | -1630.85 |
| 19 | -1632.17 |
| 20 | -1634.21 |
| 21 | -1636.51 |
